# Early detection of superspreaders by mass group pool testing can mitigate COVID-19 pandemic

**DOI:** 10.1101/2020.04.22.20076166

**Authors:** M.B. Gongalsky

## Abstract

**Background:** Most of epidemiological models applied for COVID-19 do not consider heterogeneity in infectiousness and impact of superspreaders, despite the broad viral loading distributions amongst COVID-19 positive people (1 – 10^6^ per mL). Also, mass group testing is not used regardless to existing shortage of tests. I propose new strategy for early detection of superspreaders with reasonable number of RT-PCR tests, which can dramatically mitigate development COVID-19 pandemic and even turn it endemic.

**Methods:** I used stochastic social-epidemiological SEIAR model, where S-suspected, E-exposed, I-infectious, A-admitted (confirmed COVID-19 positive, who are admitted to hospital or completely isolated), R-recovered. The model was applied to real COVID-19 dynamics in London, Moscow and New York City.

**Findings:** Viral loading data measured by RT-PCR were fitted by broad log-normal distribution, which governed high importance of superspreaders. The proposed full scale model of a metropolis shows that top 10% spreaders (100+ higher viral loading than median infector) transmit 45% of new cases. Rapid isolation of superspreaders leads to 4-8 fold mitigation of pandemic depending on applied quarantine strength and amount of currently infected people. High viral loading allows efficient group “matrix” pool testing of population focused on detection of the superspreaders requiring remarkably small amount of tests.

**Interpretation:** The model and new testing strategy may prevent thousand or millions COVID-19 deaths requiring just about 5000 daily RT-PCR test for big 12 million city such as Moscow. Though applied to COVID-19 pandemic the results are universal and can be used for other infectious heterogenous epidemics.

**Funding:** No funding

## Introduction

Coronavirus disease 2019 (COVID-19) caused by severe acute respiratory syndrome coronavirus 2 (SARS-CoV-2) occurred in Wuhan, China, in December 2019 and unprecedentedly rapidly spreaded worldwide including more than 2.5 million confirmed cases and almost 170 000 deaths at 23^th^ of April 2020 according to WHO reports. The pandemic has huge impact on quality of life, economics, because of relatively high mortality and intensive spreading, which resulted in severe lockdowns, quarantines, lack of intensive care units (ICU), overflows of national health care systems, etc. The disease is characterized by high case fatality rates (CFR) for patient over 70 years (symptomatic CFR 4-6%), reproductive number, R_0_, in the range of 1.5-3 and prevailed transmission by airborne respiratory droplets and fomites such as hands, surfaces, etc. ^1^

The spreading of COVID-19 can be mitigated by quarantine applied to majority of the population or to suspected people only. The latter one requires express and precise mass testing of the suspected people, who are usually people with symptoms or ones contacted with them. There are some diagnostic approaches for detection of viral infections including reverse transcription polymerase chain reaction (RT-PCR), enzyme-linked immunosorbent assays (ELISA) and virus isolation (VI) ^2^. New impedance-based approach is under development ^3^. The most widely used diagnostics for COVID-19 detection is RT-PCR now, because VI is fairly time-consuming and unsafe, ELISA is hardly available due to lack of targeting antibodies for SARS-CoV-2. RT-PCR usually demonstrates the highest detection rate amongst all diagnostic methods, which exceeds 80% ^2^, but it can vary depending on vendor.

The amount of RT-PCR tests is always limited (highest in Italy: 1 per 1000 of population daily ^4^), therefore optimization of testing policy is helpful. One of the polices is group testing for population with low prevalence of infection, when samples of from 2 to 64 people are mixed in a single “pool”. This allows significant reduction of tests amount, but the drawback is a decrease of sensitivity and, consequently, increase of false negative responses ^5^. More sophisticated 2D-matrix group testing was proposed for detection of COVID-19 ^6^. The testing scheme was optimized for different prevalence, but trade in between sensitivity and quantity of tests was still present.

Another important factor for spreading rate of epidemic is the heterogeneity of its infectors. COVID-19 is likely to be highly heterogeneous, because it is similar to SARS epidemic, which was also caused by coronavirus and showed 85% transmission produced by 20% of the most contagious people, also called superspreaders ^7^. Although there is no confirmed COVID-19 superspreader cases in peer-reviewed scientific literature, there are some evidences for existence of them in public media ^8^. Superspreading is present for most epidemics ^9^ and their removal from the infected population is promising. Moreover, therapeutical approaches specifically targeted on superspreaders were proposed^10^.

Another evidence for abundance of superspreaders of COVID-19 is viral loading statistics obtained by real time RT-PCR ^11–13^, which covers 6 orders of magnitude for different COVID-positive patients. Broad distributions of viral copies were observed for nasal and throat swabs and for sputum ^11^. The latter one is the former substrate to exhaled airborne droplets. Therefore broad distribution of number of virions in airborne particles was also shown for different viruses including coronaviruses and influenza ^14^. Similar broad distributions of viral copies was found for SARS ^15^. The statistics of viral loading is inherited not only for aerosol droplets, but also for fomites ^16^. Viral loading distributions are similar for both mild and severe cases of COVID-19, however severe cases show higher average loading value ^12^. The discussed distributions may follow log-normal law, which was observed for various biological systems from fruit weight of pumpkins ^17^ to bacteria population on the leafs of crops^18^. In brief, it can be explained as non-linear response of to a gaussian perturbation ^17^ or generation of pink noise as a result of bifurcation of a living system ^19^.

Computer simulations based on various mathematical models are widely used for prediction of evolution of pandemics and help to make decisions and choose appropriate governmental interventions^20^. The models can be stochastic or deterministic, and most of them are based on SEIR (S-suspected, E-exposed, I-infectious, R-recovered) approach or its modifications. Simulations can give estimations for R_0_ ^21^, predict probability of spreading of pandemic to new geographical areas ^22^, or calculate influence of isolation delay of infectious people on the spreading rate ^23^. However, the majority of the proposed models do not take into account heterogeneity of infectiousness and existence of superspreaders except rare ones ^24^.

Thus, the aim of the present article is to describe possible group pool testing strategy, which can detect superspreaders on early stages within reasonable amount of RT-PCR tests, and demonstrate the efficiency of the strategy by means of SEIR derivative model Monte Carlo simulations applied for London, Moscow and New York City as examples.

## Results and discussion

### 1. Compartments of the model

I used stochastic SEIAR compartments model (***A*** stands for admitted – see below) resolved by Monte-Carlo simulations. The model emulates behavior of ***n*** people in a city. The structure of the model was inspired by Moscow social life, however the results are also applicable for other cities, counties, other societies, etc. The program code was written in python language and provided in Suppl. Info. SEIAR model means that each citizen is presented in one of five groups (see Figure 1A):

**Figure 1.**
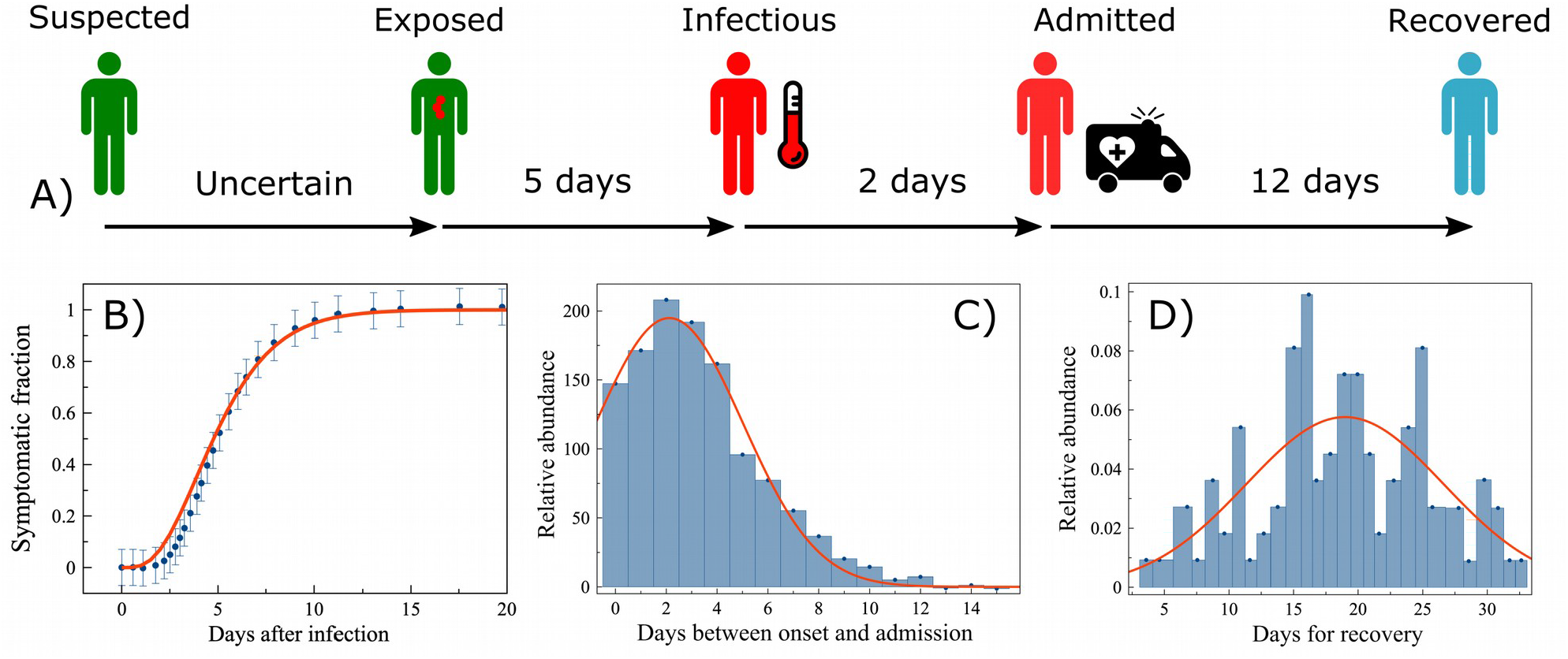
A) Model compartments sequence. S – suspected (healthy), E – exposed (partially infectious, before onset of symptoms), I – infectious (fully infectious, after onset), A – Admitted (admission to hospital or complete isolation), R – Recovered (including deceased). Average periods are shown. B) Dependence of symptoms onset fraction on period after infection (duration of incubation period). Blue dots – data from Ref. ^25^ Red curve is best fit by gamma distribution cumulative function. C) Distribution for periods between onset and admission. Blue bars – data from Ref. ^26^ Red curve is best fit gaussian. D) Recovery time distribution. Blue bars – data from Ref. ^27^ Red curve is best fit gaussian.

- S – suspected. A healthy person.
- E – exposed. A person, who already infected, but does not have symptoms yet. He can transmit infection although with smaller probability than infected person with symptoms.
- I – infectious. A person with symptoms and full probability of infection transmission.
- A – admitted. A person with symptoms, who sought medical help. The model suggests that the person got COVID-19 positive test was admitted to hospital or quarantined at home and stopped transmission of the infection.
- R – recovered. Includes both recovered and deceased. Recovered people are suggested to be immune. All of them do not influence to other compartments of the model.

Please note, that there are about 17% of asymptomatic patients according to investigation on Diamond Princess liner ^28^. The model does not take them into account for simplicity and because the infectiousness of asymptomatic people is unknown and can be negligible. However, existence of asymptomatic infectors will even support main conclusions of the article.

Each citizen has his own values of incubation period (from infections to onset of symptoms), period between onset and admission, and recovery period (see Figure 1, all curve fitting was done in MagicPlot software). All values follow standard distributions fitting respective experimental data:

- Incubation period. From exposure to onset of symptoms (E → I) – Figure 1B. Experimental data (blue circles) from Ref. ^25^ Red curve is best fit cumulative gamma distribution with shape parameter k = 4, scale parameter τ = 1.3. Average incubation period is 5 days.
- Period from onset to admission (I → A) – Figure 1C. Experimental data (blue bars) from Ref. ^26^ Red curve is best fit of gauss distribution with mean μ = 2.12 and standard deviation σ = 3.42.
- Recovery time (E → R) – Figure 1D. Experimental data (blue bars) from Ref. ^27^ Red curve is best fit of gauss distribution with μ = 19 and standard deviation σ = 9. Note, that recovery time was calculated from start of the infection not from start of the admission and the recovery may occur before the onset or admission. Average recovery time is 19 days, therefore average period between admission and recovery is 12 days as it shown in Figure 1A.

The model is stochastic, therefore it does not use the concept of serial interval ^29^, which was used for many deterministic models. Serial interval is average period between infection of a person and his first transmission. Serial interval is difficult to measure from common clinical data.

All the SEIAR population discretely changes daily in accordance with sequence presented in Figure 1A until amount of infected population (IP) (exposed+infectious+admitted) becomes 0. Initially infected people in population is chosen randomly in order to have 10-50 infected in a city.

### 2. Pattern of daily contacts

Schematic view of daily contacts is shown in Figure 2. All population of the city is split into 3 group:

**Figure 2.**
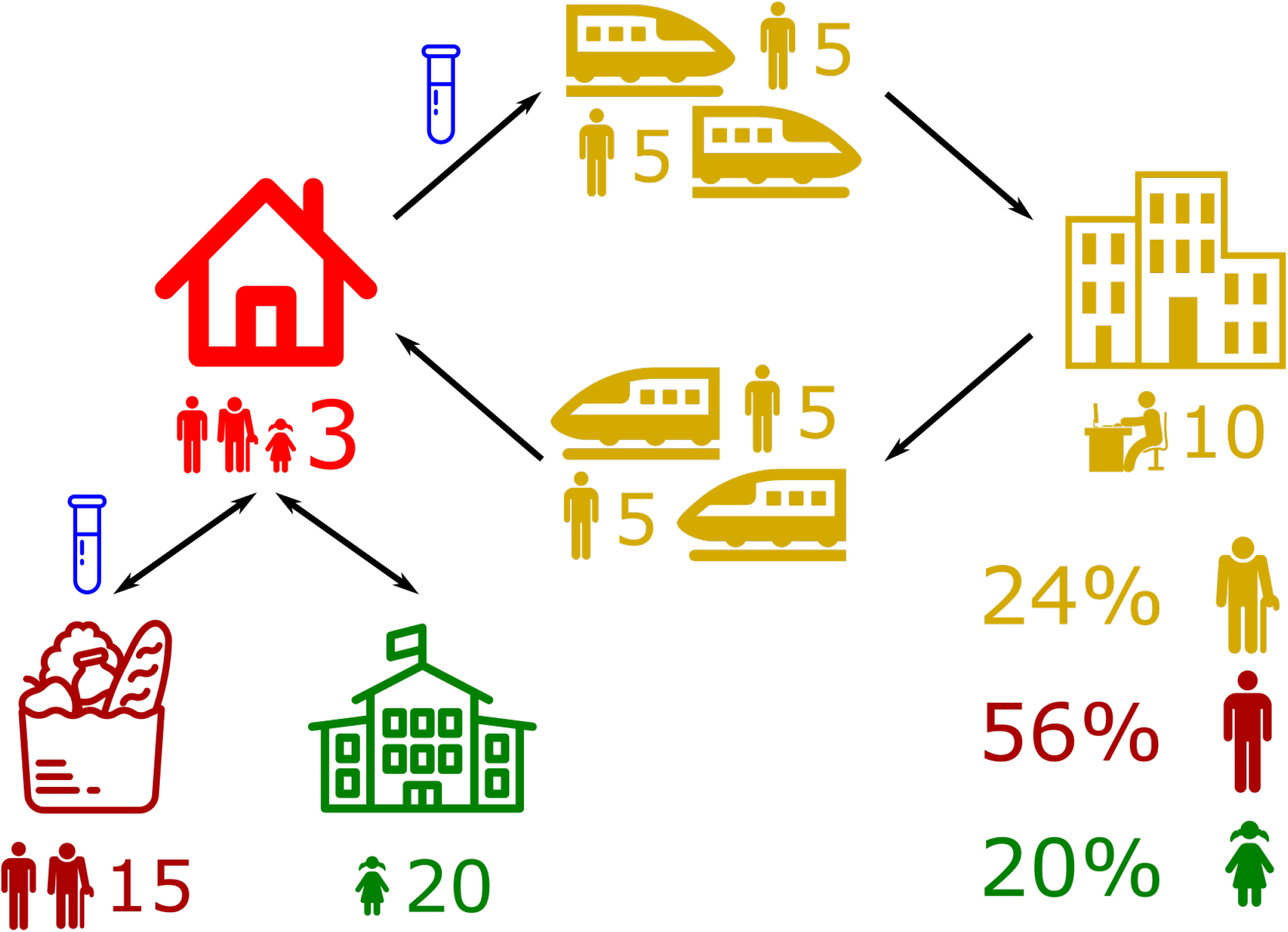
Pattern of daily contacts. Social groups: retired – men with a cane; able-bodied – featureless men, children and students – girls with pigtails. Locations: house (red), trains (2 each way - yellow), office (yellow), grocery (dark red), school (green). Pics under each location show possible attendants, numbers show sizes of groups in location. Arrows show possible daily movements. Blue test tubes show locations, where mass testing can be applied. Percentages in the corner show relative abundance of social groups in city population.

- Retired – 24%. Shown as man with a cane pics.
- Adult able-bodied – 56%. Shown as simple man pics.
- Children and students – 20%. Shown as girl with pigtails pics.

The division was made in accordance with Moscow demographic statistics. Each picture represents location of contacts, amount of contacts used in model and participating social groups. Model suggests that exactly 3 people live in each house (shown red in left top corner), all 3 groups can live, but the composition is random. There are 3 ways available every day:

- To office (shown yellow on the right of the Figure 2) connected by two subway trains each way, which is typical for Moscow. 5 people in each train represent amount of citizens, who are located relatively close to each other and may transmit the infection, despite that usually there are more people in a single coach, but they are scattered along the coach.
- To grocery (shown dark red in left bottom corner of Figure 2). It is suggested that attendance of the children is negligible. Groceries may also represent pharmacies or other shops.
- To school or university (shown green below the house). Teachers are not taken into account.

People in the same houses (housemates), same offices (co-workers), same schools or universities (classmates) are given once randomly and do not change from one day to another. On contrary, people in each train and grocery are absolutely random each time. That represents the real situation, when people have both regular and accidental contacts. Color of contact places shows relative susceptibility to quarantine (red color corresponds to independence to quarantine, dark red and yellow – moderate susceptibility, green – high susceptibility). Mass public events are out of scope of the model, because they are implied to be banned already.

Relative probabilities of the infections per day, ***P***_***pl***_, were proportionate to duration of the presence in particular place and tuned to make the model balanced in order to use all ways of infection transmission. P_pl_ were: 3 for house, 1 for office, 0.1 for each couch in subway, 0.5 for school and 0.05 for grocery. Absolute probability of a contagion in a place, ***P***_***i***_, was dependent on ***P***_***pl***_, total contagiousness of all people in the place, ***ΣP***_***j***_, and relative virulence of SARS-CoV-2, **P**_**vir**_.

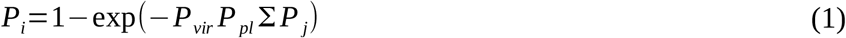

### 3. Heterogeneous infectors

The model suggests that contagiousness of a person, ***P***_***j***_, is proportionate to amount of viruses exhaled by him per minute, which is in turn proportionate to concentration of viruses in sputum or pharyngeal mucus, ***C***_***vir***_. The latter can be estimated by RT-PCR tests from sputum, throat or nasal swabs, i.e.

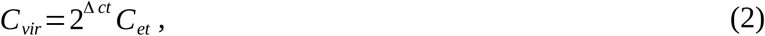

Where **Δct** is differential cycle threshold for the specimen and the etalon and **C**_**et**_ is the concentration of RNA in etalon sample.

Figure 3A shows RT-PCR cycle threshold distribution for COVID-19 positive patients shown as blue bars ^12^. Difference in **Δct** equal to 24 means that viral loading for COVID-19 patients may alter in 2^24^ = 16 million times. Error of **Δct** is about 3 ^11^. Red curve shows gaussian fit for the **Δct**, which means log-normal for viral loading. The best fit gave us: μ = 2 and σ = 3.6. Those values were used to generate distribution of simulated viral loading proportionate to contagiousness of citizens, presented in Figure 3B. The value of C_0_ corresponds to median viral loading. The distribution gives us fractions of superspreaders depending of chosen superspreaders threshold, ***St***, i.e. 10% for ×100 threshold (superspreaders are defined as people, who are 100+ times more contagious than median infectors), 5.6% for x300, 2.8% for ×1000 as it is shown in red area in Figure 3B. Data from Ref. ^12^ contain only 75 tests, but similar log-normal dependences were obtained for SARS ^15^, 778 tests of pandemic H1N1 influenza outbreak ^30^ or even for virus concentration on fomites ^16^ (see Figures S1-S3 in Suppl. Info for details).

**Figure 3.**
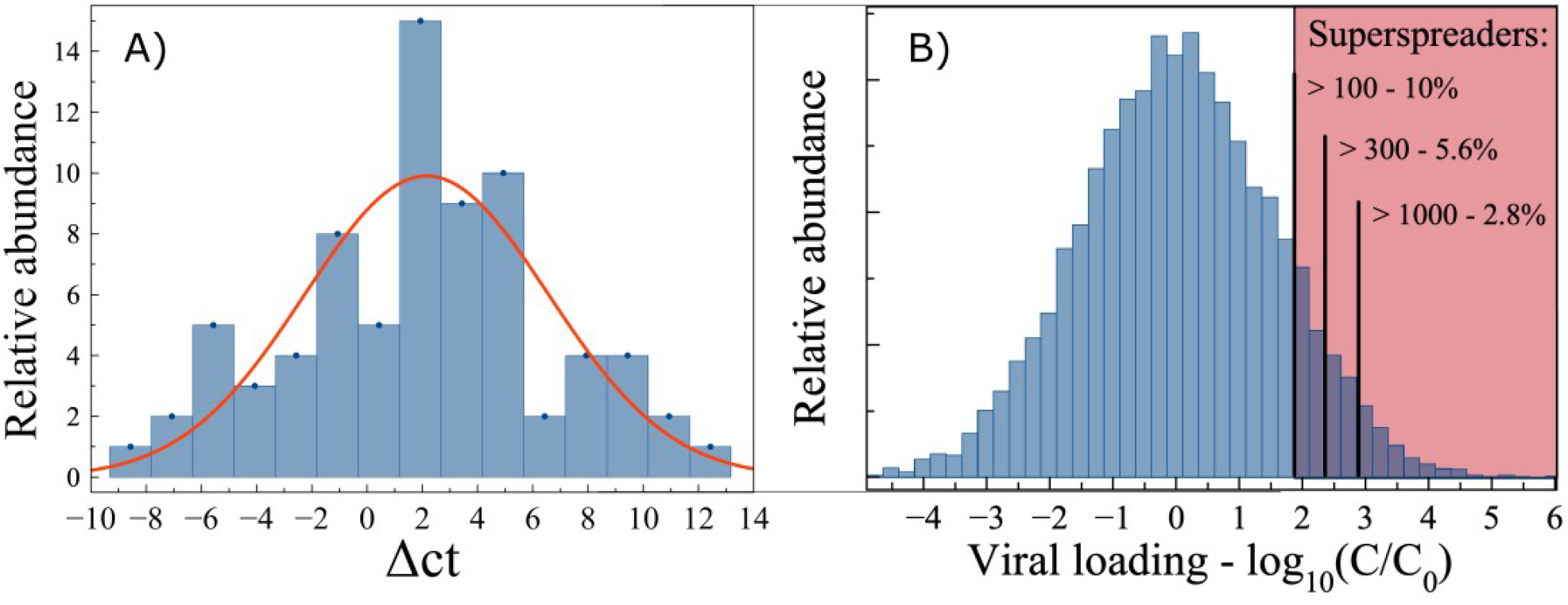
A) RT-PCR relative cycle threshold (Δct) distribution. Blue bars – data from Ref. ^12^ Red curve is best fit gaussian. B) Random generated log-normal viral loading distribution (blue bars). Red area corresponds to superspreaders. Different superspreaders thresholds are shown as black bars with corresponding prevalence.

### 4. Options: quarantines and mass testing

The model takes quarantine into account. The efficiency of quarantine for different locations and social groups is shown in Figure 2 as color legend from red to green. Contagions in houses are not affected by quarantines. Schools and universities are closed in any quarantine. Offices, trains and groceries are partially affected by quarantines. The model assumes that amount of workers or customers are dropped by the quarantine factors, ***Q***_***of***_, and ***Q***_***gr***_, correspondingly, after its beginning. Office quarantine affects certain subgroup of able-bodied citizens, who start to work remotely and do not comute to office. Other workers (like policemen, e.g.) are not affected by quarantine. Grocery quarantine affects all able-bodied and retired people, i.e. they shop less often, sometimes prefer delivery services, so their average attendance to groceries reduced, but there is no division on stable subgroups. The Q factors can be changed several times during evolution of pandemic. I used the following relationship:

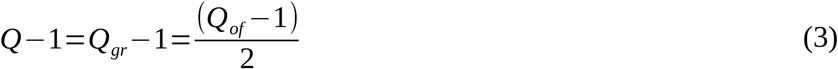

Mass testing is another option of the program and it can be applied for all customers of public transport and groceries as it marked as blue test tubes in Figure 2. The screening detects only superspreaders with given threshold and put them in complete isolation (programmed as admission) after obtaining test results (2 days). Mass testing can be switched on a particular day.

### 5. Outbreak prognosis for London, Moscow and New York City

Typical curves for all 5 SEIAR compartments are shown in Figure 4A. The sum of E, I and A was used as infected population (IP). Simulation of pandemic dynamics in London, Moscow and New York City (NYC) is shown in Figure 4B. Arrows point to quarantine interventions with corresponding Q factors. Times of IP doubling are shown above the simulated curves for all cities.

**Figure 4.**
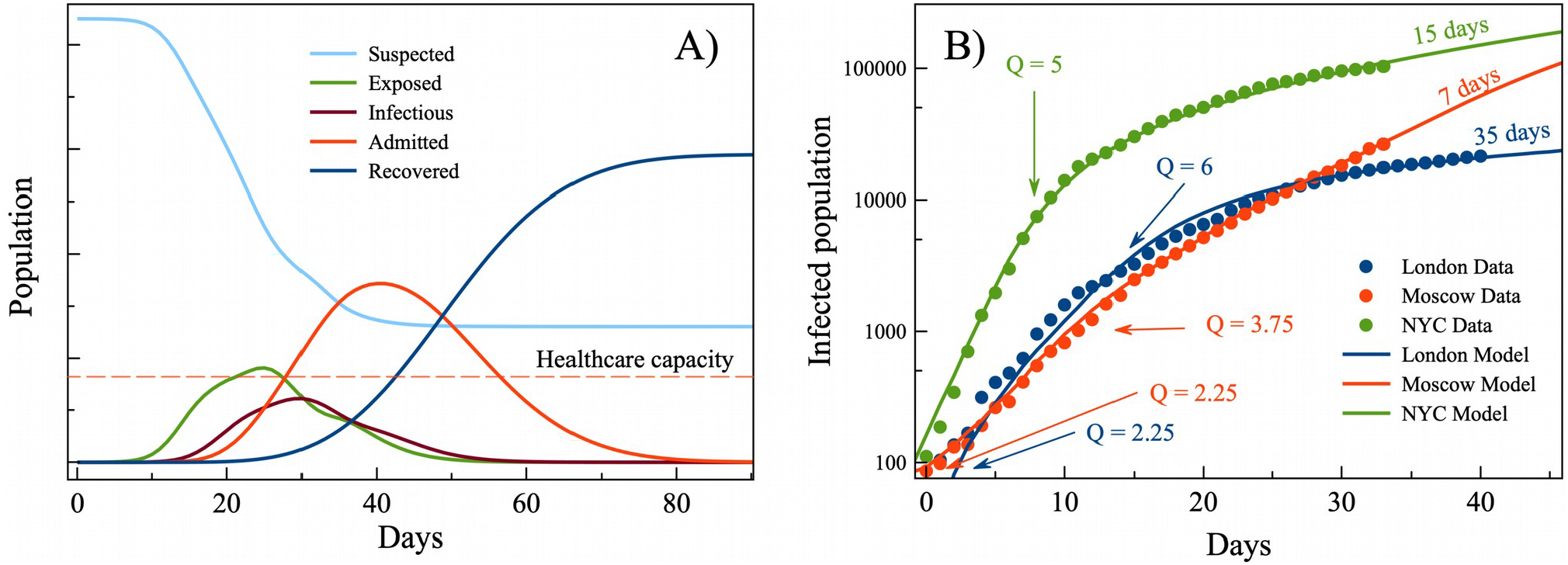
A) Typical SEIAR simulation curves. B) Official data and model simulation of COVID-19 outbreaks in London (blue), Moscow (red) and New York City (green). Arrows show introduction of quarantines with corresponding Q factors. Doubling times are shown above the curves.

Figure 5 shows COVID-19 developments in three cities without testing (blue curves), and with testing and isolation of superspreaders with St = 100 (green), 300 (yellow), 1000 (orange). Public data of confirmed cases are shown as black dots. Hospital beds and intensive care units (ICU) capacities are shown as horizontal dashed lines (see Suppl. Info for details). Rapid isolation of superspreaders leads to substantial mitigation of pandemic for both total and peak IP (shown on bar chart in Figure 5, see numerical data for Figure 5 in Suppl. Info.). For cities with strong quarantine (London and NYC) this testing strategy with St = 100 strongly reduces amount of total IP (4.5-4.8 fold decrease is predicted), while for Moscow with weak quarantine more prominent is reduction of peak IP (8-fold).

**Figure 5.**
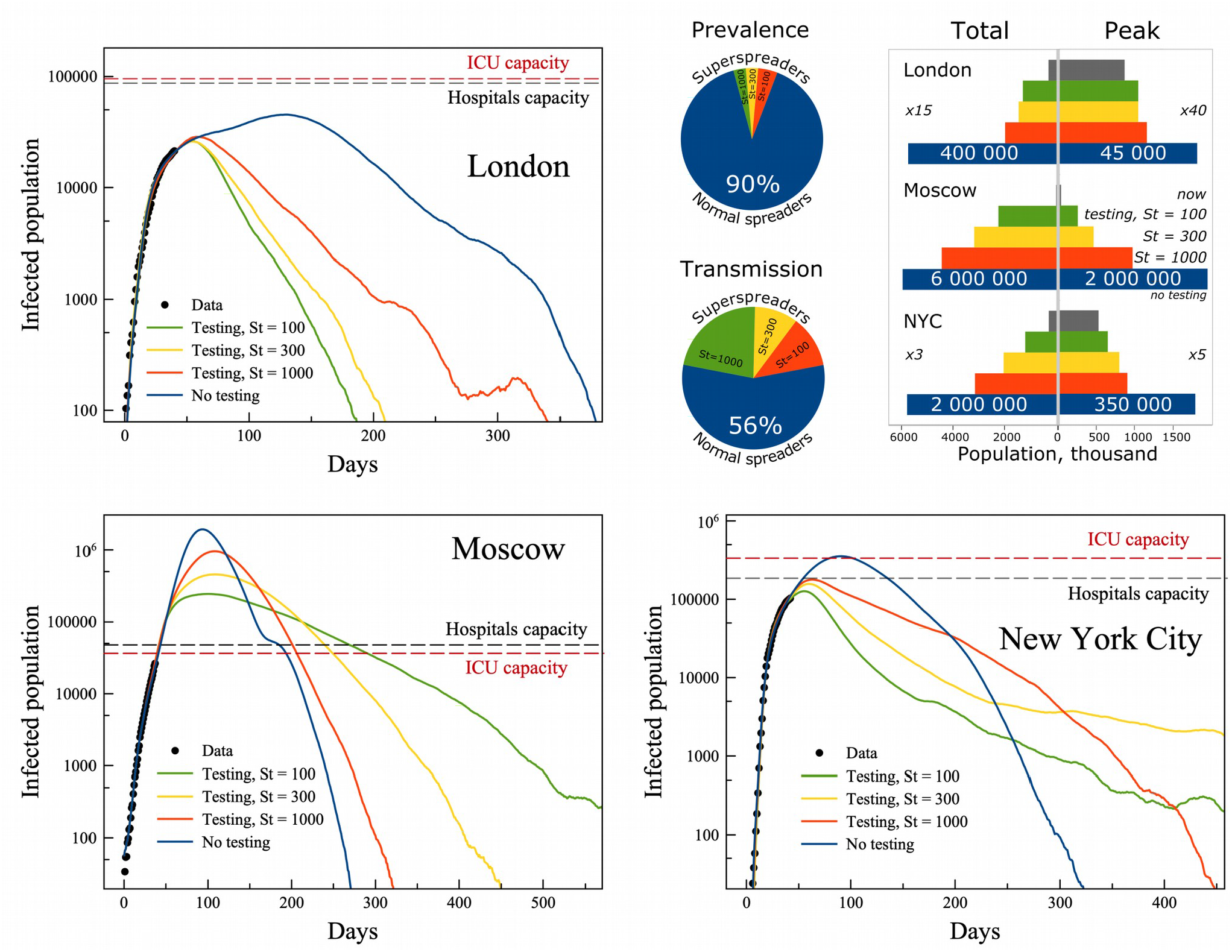
Simulated COVID-19 pandemic curves for London, Moscow and New York City without mass testing (blue), and with mass testing with different superspreaders threshold, **St**, 100 (green), 300 (yellow) and 1000 (orange). Official data are shown as black circles. Pie charts show prevalence of superspreaders between different **St** ranges and their impact on transmission. Bar charts show total and peak infected population values for all cities and testing options.

Choice of St is very important for practical implementation of mass testing strategy, because it is a trade in between difficulty and efficiency. Higher **St** corresponds to lower amount of daily required RT-PCR tests, but it fails to detect superspreaders below **St**. Prevalence of superspreaders between different **St** values as well as corresponding effect of COVID-19 transmission by them are shown on pie charts in Figure 5. Note, that regardless to **St** value, throat or/and nasal swabs must be taken from all people attending offices and groceries, which is also a challenge for metropolises. Low **St** is less important for London, where both relatively low initial IP and strong quarantine, i.e. it is 10% difference for peak IP and 50% for total IP between **St** = 100 and 1000. Differences increase for total IP with increase of initially IP and reaches 160% for NYC, while low **Q** in Moscow gives 300% reduction for peak IP. The last one is crucially important, because predicted 2 million COVID-19 patients in Moscow is far over its healthcare capacity, which can result in about 0.5 million excessive deaths. Nevertheless, the prognosis demonstrates catastrophic scenario for Moscow, therefore strong quarantine such as one used in London or NYC is highly recommended regardless of possible new testing strategy application.

### 6. Mass testing strategy

Superspreaders isolation based mitigation requires mass testing strategy for detection of them. The simplest strategy is make one test for person, but it requires about 2 million test daily, which is obviously impossible to do. However, daily tests requirements can be reduced at least by the factor of 500, if smart matrix group pool testing scheme used (see Figure 6). Note, that this scheme is very efficient for detection of superspreaders, but it misses all other infected people. The exact amount of tests depends on the superspreaders threshold **St**. If it is assumed that viral loading for median spreader is 10 times higher than sensitivity of RT-PCR test (which is true for most used test-systems), then test will give positive result for a superspreader mixed into a pool with 10*S_t_ = 1000 for St = 100 and even more for higher thresholds. But matrix testing requires 2 tests for each specimen, i.e. one in a row and one in a column. Then all specimens in the intersections (highlighted blue) can be tested separately without mixing. This will provide extremely low false positive result ratio, because viral loading is huge in positive tests. In case of low prevalence the additional amount of separate tests will be much lower than 500. Thus, that strategy allows to detect superspreaders by perform 5000 tests daily for megapolices such as Moscow, New York City and London.

**Figure 6.**
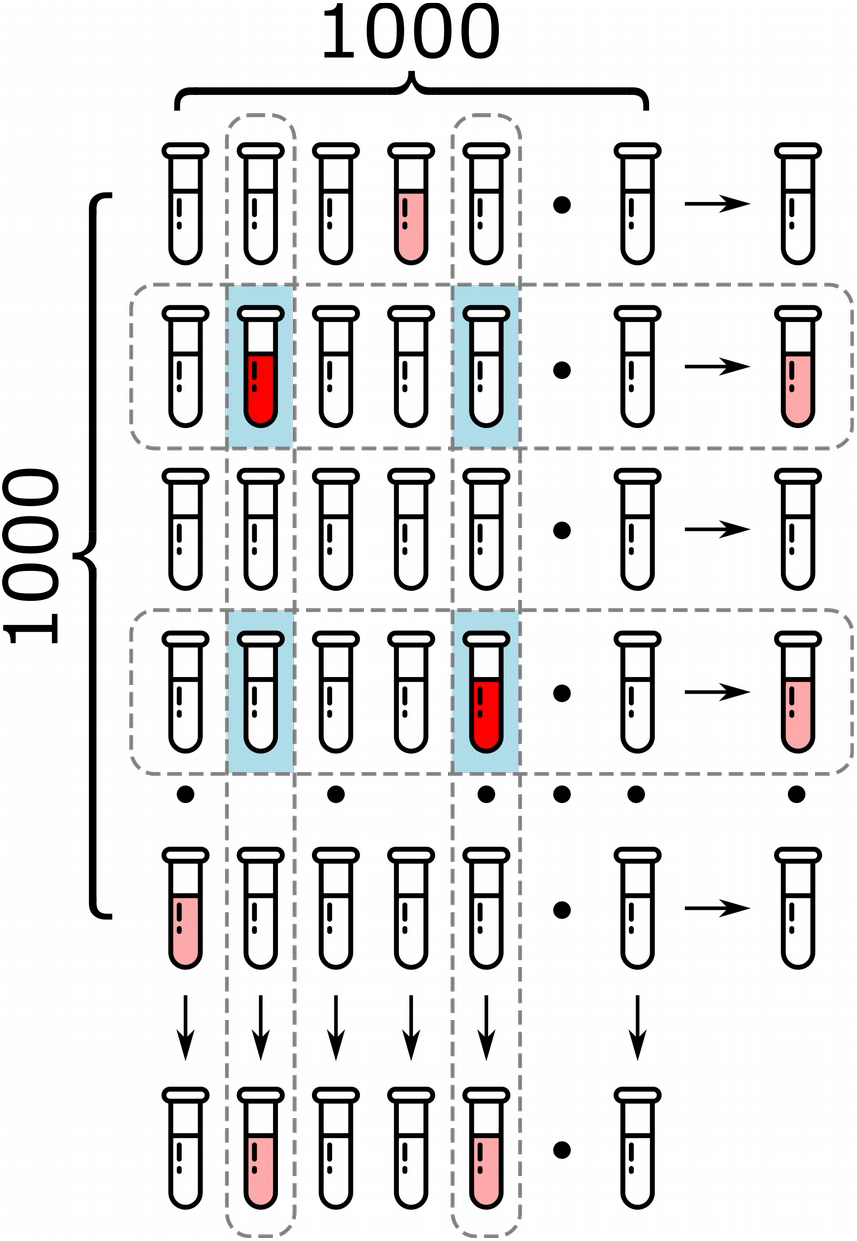
Schematic view of matrix group pool testing strategy. Specimens are mixed both in rows and columns. Red test tubes show superspreaders. Light red test tubes show normal spreaders and mixtures with superspreaders. While test tubes are negative. Light blue background highlights specimens in the intersections, which are require subsequent separate testing.

## Conclusions

Thus, the proposed stochastic SEIAR-model for COVID-19 pandemic demonstrated crucial importance of superspreaders, who are people with SERS-CoV-2 viral loading at least 100 exceeding median value. Superspreaders with 10% prevalence amongst infected people transmit 45% cases of COVID-19, therefore their rapid isolation can significantly mitigate pandemic and save thousands of people, as it was shown for London, Moscow and New York City. The isolation can be performed via mass matrix group pool strategy, applied for all people attended to offices and groceries. This strategy requires reasonable amount of RT-PCR tests about 5000 per day. The obtained results can be also applied to other cities and countries and used not only for COVID-19 pandemic, but for other infectious diseases with high heterogeneity of spreaders.

## Data Availability

All data are available.

## References

1 Wu JT, Leung K, Bushman M, et al. Estimating clinical severity of COVID-19 from the transmission dynamics in Wuhan, China. Nat Med 2020; 26: 506–10.

2 Steininger C, Kundi M, Aberle SW, Aberle JH, Popow-Kraupp T. Effectiveness of Reverse Transcription-PCR, Virus Isolation, and Enzyme-Linked Immunosorbent Assay for Diagnosis of Influenza A Virus Infection in Different Age Groups. Journal of Clinical Microbiology 2002; 40: 2051–6.

3 Gongalsky MB, Tsurikova UA, Samsonova JV, et al. Double etched porous silicon nanowire arrays for impedance sensing of influenza viruses. Results in Materials 2020; 6: 100084.

4 COVID-19 pandemic. The charts of our World in Data. https://ourworldindata.org/the-covid-19-pandemic-slide-deck.

5 Yelin I, Aharony N, Shaer-Tamar E, et al. Evaluation of COVID-19 RT-qPCR test in multi-sample pools. Infectious Diseases (except HIV/AIDS), 2020 DOI:10.1101/2020.03.26.20039438.

6 Sinnott-Armstrong N, Klein D, Hickey B. Evaluation of Group Testing for SARS-CoV-2 RNA. Infectious Diseases (except HIV/AIDS), 2020 DOI:10.1101/2020.03.27.20043968.

7 Lloyd-Smith JO, Schreiber SJ, Kopp PE, Getz WM. Superspreading and the effect of individual variation on disease emergence. Nature 2005; 438: 355–9.

8 Lanese N. ‘Superspreader’ in South Korea infects nearly 40 people with coronavirus. https://www.livescience.com/coronavirus-superspreader-south-korea-church.html.

9 Galvani AP, May RM. Dimensions of superspreading. Nature 2005; 438: 293–5.

10 Metzger VT, Lloyd-Smith JO, Weinberger LS. Autonomous Targeting of Infectious Superspreaders Using Engineered Transmissible Therapies. PLoS Comput Biol 2011; 7: e1002015.

11 Yu F, Yan L, Wang N, et al. Quantitative Detection and Viral Load Analysis of SARS-CoV-2 in Infected Patients. Clinical Infectious Diseases 2020; : ciaa345.

12 Liu Y, Yan L-M, Wan L, et al. Viral dynamics in mild and severe cases of COVID-19. The Lancet Infectious Diseases 2020; : S1473309920302322.

13 Wang W, Xu Y, Gao R, et al. Detection of SARS-CoV-2 in Different Types of Clinical Specimens. JAMA 2020; published online March 11. DOI:10.1001/jama.2020.3786.

14 Leung NHL, Chu DKW, Shiu EYC, et al. Respiratory virus shedding in exhaled breath and efffcacy of face masks. Nat Med 2020; published online April 3. DOI:10.1038/s41591-020-0843-2.

15 Chu C-M, Cheng VCC, Hung IFN, et al. Viral Load Distribution in SARS Outbreak. Emerg Infect Dis 2005; 11: 1882–6.

16 Zhang N, Li Y. Transmission of Influenza A in a Student Offfce Based on Realistic Person-to-Person Contact and Surface Touch Behaviour. IJERPH 2018; 15: 1699.

17 Koch AL. The logarithm in biology. Journal of Theoretical Biology 1969; 23: 251–68.

18 Hirano SS, Nordheim EV, Arny DC, Upper CD. Lognormal Distribution of Epiphytic Bacterial Populations on Leaf Surfaces. Applied and Environmental Microbiology 1982; 44: 695–700.

19 Szendro P, Vincze G, Szasz A. BIO-RESPONSE TO WHITE NOISE EXCITATION. Electro- and Magnetobiology 2001; 20: 215–29.

20 Flaxman S, Mishra S, Gandy A, et al. Report 13: Estimating the number of infections and the impact of non-pharmaceutical interventions on COVID-19 in 11 European countries. 2020; : 35.

21 Liu Y, Gayle AA, Wilder-Smith A, Rocklöv J. The reproductive number of COVID-19 is higher compared to SARS coronavirus. Journal of Travel Medicine 2020; 27: taaa021.

22 Kucharski AJ, Russell TW, Diamond C, et al. Early dynamics of transmission and control of COVID-19: a mathematical modelling study. The Lancet Infectious Diseases 2020; : S1473309920301444.

23 Hellewell J, Abbott S, Gimma A, et al. Feasibility of controlling COVID-19 outbreaks by isolation of cases and contacts. The Lancet Global Health 2020; 8: e488–96.

24 Fujie R, Odagaki T. Effects of superspreaders in spread of epidemic. Physica A: Statistical Mechanics and its Applications 2007; 374: 843–52.

25 Lauer SA, Grantz KH, Bi Q, et al. The Incubation Period of Coronavirus Disease 2019 (COVID-19) From Publicly Reported Confirmed Cases: Estimation and Application. Ann Intern Med 2020; published online March 10. DOI:10.7326/M20-0504.

26 Donnelly CA, Ghani AC, Leung GM, et al. Epidemiological determinants of spread of causal agent of severe acute respiratory syndrome in Hong Kong. The Lancet 2003; 361: 1761–6.

27 Verity R, Okell LC, Dorigatti I, et al. Estimates of the severity of COVID-19 disease. Epidemiology, 2020 DOI:10.1101/2020.03.09.20033357.

28 Mizumoto K, Kagaya K, Zarebski A, Chowell G. Estimating the asymptomatic proportion of coronavirus disease 2019 (COVID-19) cases on board the Diamond Princess cruise ship, Yokohama, Japan, 2020. Eurosurveillance 2020; 25. DOI:10.2807/1560-7917.ES.2020.25.10.2000180.

29 Nishiura H, Linton NM, Akhmetzhanov AR. Serial interval of novel coronavirus (COVID-19) infections. International Journal of Infectious Diseases 2020; 93: 284–6.

30 Yang J-R, Lo J, Ho Y-L, Wu H-S, Liu M-T. Pandemic H1N1 and seasonal H3N2 influenza infection in the human population show different distributions of viral loads, which substantially affect the performance of rapid influenza tests. Virus Research 2011; 155: 163–7.

